# Temporal and interaction dynamics of dengue cases, entomological and meteorological variables in Melaka, Malaysia: A multivariate time series analysis

**DOI:** 10.1101/2024.08.30.24312846

**Authors:** Shazelin Alipitchay, Muhammad Aswad Alias, Sharifah Nur Shahirah Syed Abdul Hamid, Rabizah Hamzah, Norain Mansor, Nurulhusna Ab. Hamid, Hidayatulfathi Othman

**Author notes:** Corresponding author (SA). These authors contributed equally to this work (Shazelin Alipitchay, Muhammad Aswad Alias). These authors also contributed equally to this work (Sharifah Nur Shahirah Syed Abdul Hamid, Rabizah Hamzah).

## Abstract

The interaction between dengue cases, entomological and meteorological variables has remained intricate for decades. Validated facts are important to form robust decision making with the adoption of safer and sustainable efforts. This study aims to elucidate the relationship between the variables in the long run and short-term dynamic focusing in Melaka, Malaysia, in an attempt to improve the understanding of the variables and their temporal associations. This study quantifies the variables on their temporal associations, potential time lags, and dynamic interplays between all the variable data sets. The research applies a Johansen Cointegration Test and Vector Error Correction Model to validate long term run and examine short-term deviations among dengue cases, temperature, ovitrap and sticky ovitrap data from 2020-2022. Empirical findings prove that temperature, sticky ovitrap index (SOI) and ovitrap index (OI) has a significant and unique long-run equilibrium relationship with dengue cases. The short-term equilibrium results display a robust causality between variables. The model fit elucidates 74.2% of the dynamics. The VECM model provides an excellent trade-off between goodness of fit and complexity in describing the variables examined. Previous dengue occurrences predicted a surge of new dengue cases while preserving the cyclical pattern. The model predicts the utility and efficacy of sticky ovitraps. It also validates ovitrap use as a surveillance tool and offers substantiation of the influence of temperature on the progression of dengue cases.

## Introduction

According to the World Health Organization, dengue fever is currently regarded as the most critical mosquito-borne viral disease worldwide. Furthermore, it is the most rapidly spreading, with a 30-fold surge in worldwide occurrence within the last five decades. The Western Pacific and Southeast Asia regions collectively represent 75% of the worldwide dengue illness burden [1]. The relationship among Dengue virus (Den V), Aedes mosquitoes, humans as the principal host, and the environment is intricate. Due to the absence of viable therapies and vaccinations, disease surveillance and vector control populations remain the primary components of the current dengue prevention program [2]. A new paradigm has been proposed, focusing on rapid, sensitive, and intensive control methods that incorporate intensive vector surveillance to curb and reduce the Aedes population [3]. In order to establish a more refined and practical strategic control method, it is essential to comprehend and investigate the critical variables—host, environment, and entomological relations—in order to facilitate more expeditious and robust decision-making.

Evidence on the dynamic interaction of these variables is still insufficient worldwide. Many studies use correlation and comparison analysis, which leads to inconclusive findings and insufficiently informed decision-making. To fully understand the underlying trends or systemic patterns over time, it is essential to use time series analysis between the three main variables. This will enable us to examine the relationships between each variable, draw conclusions, and propose necessary changes that should be made to control and prevent dengue. It is important to note that time-series analysis isn’t just about predicting the future; instead, it’s about understanding the past. Studies that utilize entomological data to forecast dengue cases are still scarce [4,5] and provide insufficient insight and conclusion. The entomological studies measure the infestation level of aedes using a proximal variable such as number of larvae. The studies are unable to provide sufficient data to explain the dynamic interaction and accurately forecast the model. The reasons may be based on the multifaceted dynamics and interactions of disease transmission, which involve complex aedes behavioural and bionomics, as well as the disease manifestation within the host system.

The effects of meteorological variables on dengue cases are yet to be explored globally with the use of time series. Only in recent times have studies begun to investigate the influence of climatic conditions on the occurrence of dengue cases. A study in Malaysia used a multilinear regression analysis to investigate the relationship between meteorological variables and dengue cases. The findings showed a positive correlation between rainfall and temperature with weekly dengue cases [6]. Additionally, the study looked at the strongest correlation between the ground-based stations and the satellite-based Earth. It found that temperature, followed by rainfall and wind speed, is a useful meteorological variable for predicting dengue in future studies. A study conducted in India utilised different regression model to determine the impact of meteorological parameters on dengue occurrences. The results revealed that humidity and mean maximum temperature are the primary factors influencing dengue incidences, with significant positive and negative associations respectively [7]. Relative humidity was found to be the most accurate indicator of increasing dengue cases in a study in Bangkok [8].

Two experimental studies on dengue cases were conducted in Malaysia to create a forecasting model by analyzing environmental, entomological, and epidemiological data series. Both studies highlighted the forecasting element. The first study showed only two significant variable data sets: the relation between entomological and environmental variables with the use of the autoregressive distributed lag (ADL) model [9]. The ovitraps data was used as the entomological variable. The previous week’s rainfall played a significant role in increasing the mosquito population, followed by maximum humidity and temperature. The second study was implemented in dengue-prone areas with the use of ovitraps and several mobile weather stations comprising rain gauges, temperature, and humidity data loggers located in the areas [10]. This is a follow-up study to address the cofounding factors and improve forecast accuracy. Significant findings were established among the three variables, with a forecasting accuracy of 85% for the specific area. Another study with the use of data from the two previous studies mentioned, proposed the installation of mobile rain gauges inside each dengue risk area to be integrated into the early warning system in order to generate a more precise prediction data [11]. This current study incorporates three variables: epidemiological, entomological, and meteorological data (temperature). Two sets of serial entomological data are being analyzed to measure the aedes infestation level: ovitraps and sticky traps. This research attempts to elucidate the relationship between the variables in the long run and short term with the use of multivariate time series analysis. From this multivariate time series analysis, this study hopes to achieve and quantify the variables on their temporal associations, potential time lags, and dynamic interplays between all the variable data sets. It can offer insights into the cointegration of all variables, improve the understanding of the variables that have impact on the incidence of dengue cases, and guide specific targeted interventions and control measures in the area. The temperature variable was selected as it has an excellent correlation between the ground-based stations and equivalent satellite-based Earth Observation (EO) data, making it an outstanding estimator of meteorological variables.

## Materials and methods

### Study area

This study is in Melaka Tengah, a high-density population with 1,673 people per square kilometer. The state consists of three districts: Melaka Tengah, Alor Gajah, and Jasin. The land covers an area of 1664 km^2^. It has a population of 985700 people. The state is situated in the southern part of Peninsular Malaysia. As of July 7, 2008, Malacca City, the capital of the state, has been recognized as a UNESCO World Heritage Site, attracting numerous tourists from around the globe. The city experiences an equatorial climate with high temperatures and humidity all year round. It receives heavy rainfall, especially between September and November. In the past five years, daytime temperatures have ranged from 31 to 33 °C (88 to 91 °F), while nighttime temperatures have averaged around 23 °C (73 °F).

### Epidemiological data

Melaka recorded a total of 2843 dengue cases in 2020 with an incidence rate of 295 per 100,000 population, 611 dengue cases in 2021 with an incidence rate of 62.8 per 100,000 population, followed by a total of 665 dengue cases in 2022 with an incidence rate of 67 per 100,000 population [12]. Melaka Tengah accounted for the majority of reported dengue cases (70%). In Malaysia, dengue cases are reported as soon as a diagnosis is made and entered into an electronic notification web-based system. This system is then directly linked to a second web-based system called e-Dengue, which records all the profiles and locations of dengue cases. e-Dengue consists of data that has been validated epidemiologically and geocoded. The system also records control activity information. Thus, it is specifically designed to generate official daily report instantly. The data set on dengue cases included all registered cases that occurred between epidemiological week 1, 2020, and epidemiological week 52, 2022, spanning a total of 157 consecutive weeks.

### Entomological data

Two types of Aedes traps were employed: the conventional ovitrap and the autocidal trap. They have been used as a standard entomological tool to survey the mosquito population. An ovitrap is a cylindrical plastic container with opaque black sides, which has a diameter of 7 cm and a height of 9 cm. A hardboard oviposition paddle of 10 cm × 3.0 cm × 2.5 cm is inserted into each ovitrap, with the rough surface facing upwards. The ovitrap was filled with tap water up to a height of 5.5 cm. A total of 60 ovitraps were installed within premises, semi-indoor, and outdoor. They are placed on weekly basis at Station 1, located in Batu Berendam ecosystem, a public residential area. These containers were positioned there subsequent to acquiring consent from the homeowner.

The second trap, known as the sticky trap or MyMAT (Malaysian Mosquito Autocidal Trap), is comprised of a 700-ml black plastic cylinder container. It is filled with approximately 600 ml of dechlorinated water and BTI (Bacillus thuringiensis) larvicide. An adhesive plastic strip, commonly known as a sticky card, was inserted into the container to capture the gravid adult mosquito during oviposition. The dechlorinated water, together with BTI and sticky cards, were replaced every two weeks. Then, the adult mosquitoes were extracted from the adhesive card using forceps, then identified using a 20X magnifying glass. The relevant data was subsequently recorded in the field during the trap inspections. A total of 200 sticky traps were strategically positioned similarly to ovitraps and were deployed at Station 2 within the Sungai Udang ecosystem, which is another public residential area. Both the ovitrap index and the sticky trap index can serve as estimators for the size of the Aedes population, the size of the larvae, and the size of the adults, respectively [13–15]. Sticky traps that capture adult female Aedes mosquitoes provide valuable insights into the likelihood of dengue transmission during an ongoing outbreak.

### Meteorological data

Temperature data set from 2020 up to 2022 was retrieved from Meteorological Department.

### Data processing

R software 3.2.0 version is used to analyzed the time series data. The dataset comprises records of dengue cases (onset), ovitrap index, sticky trap index, and temperature observations spanning from 2020 to 2022. Each of the four data sets was graphed on its own panel, with a shared horizontal axis representing time in serial biweekly intervals.

### Data process flow

#### Stationarity testing and optimal lag estimation

The Augmented Dickey-Fuller test was employed to assess the stationarity of each dataset and time series. The null hypothesis, that a unit root is present, would be rejected for a series if its p-value fell below the chosen significance level of 5%, indicating stationarity. The non-stationary series were made stationary by differencing the series and tested for stationarity until they achieved stationarity. It entails the need to use a specific model, ensuring that subsequent analysis yields valid and reliable results for forecasting or understanding the dynamics between these variables. The optimal lag was selected based on the Akaike information criterion, AIC [16]. A p-1 lag was applied in estimation, referring to the lag order of the error correction term. VECM affords flexibility in terms of identifying both short-term dynamics and long-run equilibrium relationships between integrated variables. Careful attention to stationarity testing and cointegration analysis can thus help ensure the appropriateness and meaningfulness of any econometric models employed and conclusions drawn.

#### Cointegration testing

The cointegration of the variables was assessed using the Johansen cointegration test of the eigenvalue approach. The test is to detect multiple cointegration time series and is appropriate for multivariate analysis time series [17]. The null hypothesis of rank = 0 means that there is no cointegration at all.

#### Model estimation

The estimation of the model used and selected is done by looking into a series of cointegrations. If the series are not cointegrated, we can estimate the model via the vector autoregressive (VAR) function. If the series are cointegrated, the Vector Error Correction Model (VECM) is considered. This differs from the vector autoregressive (VAR) model in terms of the error correction term. VECM is a multivariate time series analysis that will compute the effect of how the growth rate of a variable changes if one of the variables departs from its equilibrium value. Hence, it allows both short-run and long-run coefficients, providing insights into equilibrium relationships and deviations from that equilibrium. VECM is able to capture the causal effect and determine the short- and long-term runs. Therefore, it is plausible and relevant in explaining the case-occurrence pathway.

#### Forecasting evaluation

Forecasting model based on VECM was captured with the use of comparative analysis to assess the forecasted biweekly onset values compared against biweekly average actual values. It is observed over a 12-week period, commencing from January 2023 until March 2023. Three primary statistical metrics, Mean Squared Error (MSE), Root Mean Square Error (RMSE) and Mean Absolute Percentage Error (MAPE) were employed to quantify the deviations between the VECM forecasts from the actual observed values.

## Results

### Time plot data series

The time series analysis of four distinct variables is depicted in the diagram (Figure 1): case counts (dengue onsets), temperature (tem), ovitrap index (oi), and sticky ovitrap index (soi). A common horizontal axis that represents time is shared by each variable, which is plotted in a distinct panel. The time plot is translated into biweekly serial data, resulting in 78 observations.

**Figure 1.**
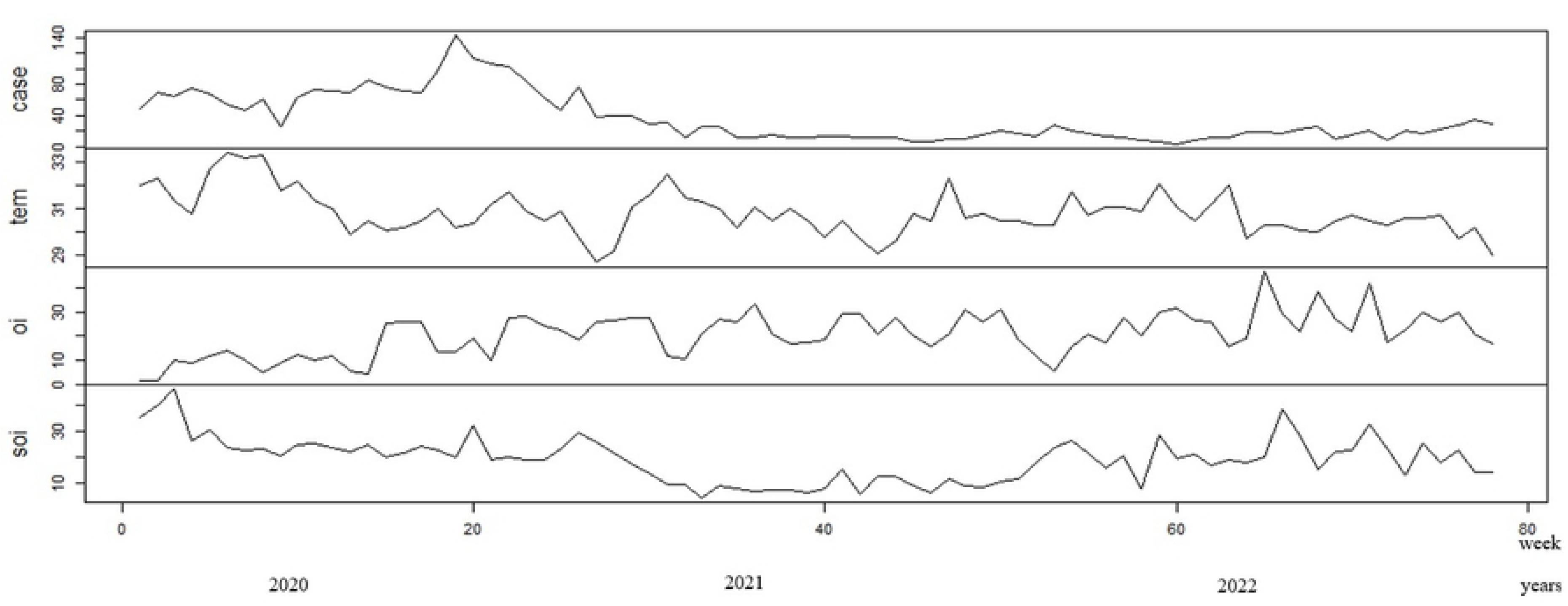
Four distinct panel of line graphs with case counts (dengue onsets), temperature (tem), ovitrap index (oi), and sticky ovitrap index (soi) readings on y-axis plotted against time in week from 0 up to 80 weeks (year from 2020 till 2022) on x axis, extracted from R.

The case count exhibits significant fluctuation in the early stages but becomes rather consistent in later periods. In 2020, the number of dengue cases reached its greatest level of 1948 cases, with an incidence rate of 332.1 per 100,000 people. In comparison, there were 478 cases with an incidence rate of 80.4 in 2021, and 444 cases with an incidence rate of 73.8 in 2022. The temperature shows some fluctuation with no clear seasonal pattern discernible from this graph with minimum average temp of 28.7°C up to maximum average temperature of 33.4°C. The ovitrap index exhibits prominent deviations with peaks and troughs, while the sticky ovitrap index for adult mosquitoes demonstrates significant volatility with occasional spikes, potentially suggesting sudden increases in mosquito populations at specific intervals.

The number of cases varies in the beginning but reach to a fairly stable level later. With 1948 cases, the number of dengue cases peaked in 2020, with an incidence rate of 332.1 per 100,000. In contrast, the incidence rate of 478 cases in 2021 was 80.4, while the incidence rate of 444 cases in 2022 was 73.8. This graph also indicates that there is no obvious seasonal pattern in the temperature data series, with a minimum average temperature of 28.7°C and a maximum average temperature of 33.4°C. The sticky ovitrap index for adult mosquitoes shows considerable volatility with sporadic spikes, which may indicate abrupt increases in mosquito populations at particular periods. The ovitrap index shows notable deviations with peaks and troughs.

### Augmented Dickey Fuller, ADF unit root testing

The Augmented Dickey-Fuller (ADF) test successfully identified the existence of three non-stationarity data series, which were then transformed into stationary series using first difference. Guided by AIC, a lag length of 10 was determined to be optimal, providing a balance between model complexity and model fitting. This lag order is used for estimating Johansen cointegration and VECM tests.

### Cointegration Analysis

#### Cointegration trace test statistics

A 10-lag vector autoregression model was utilized to conduct the Johansen cointegration trace test on the variables. The findings show that there is one cointegrating equation with a significant level, indicating a long-term equilibrium relationship between the variables. Table 1 presents the eigenvalues and trace test statistics for four hypotheses. These hypotheses involve ranks of ≤3, ≤2, ≤1, and =0. The greatest value obtained from these tests is 0.477. The following column displays the trace test statistics alongside crucial values corresponding to specific confidence levels: 10%, 5%, and 1%. Applying the conventional method of using a 5% critical value, with ‘r’ denoting the rank and ‘r = 0’, the test statistics of 78.2 is greater than 53.1. Subsequently, null hypotheses that propose ‘r > 0’, which implies the existence of cointegration was rejected. When the value of r is less than or equal to 1, the test statistics of 33.44 is less than 34.9, the null hypotheses was not rejected. Ultimately, there is at most one cointegration relationship that exists at a significant level, elucidating a long-term state of balance among the variables.

**Table 1.**
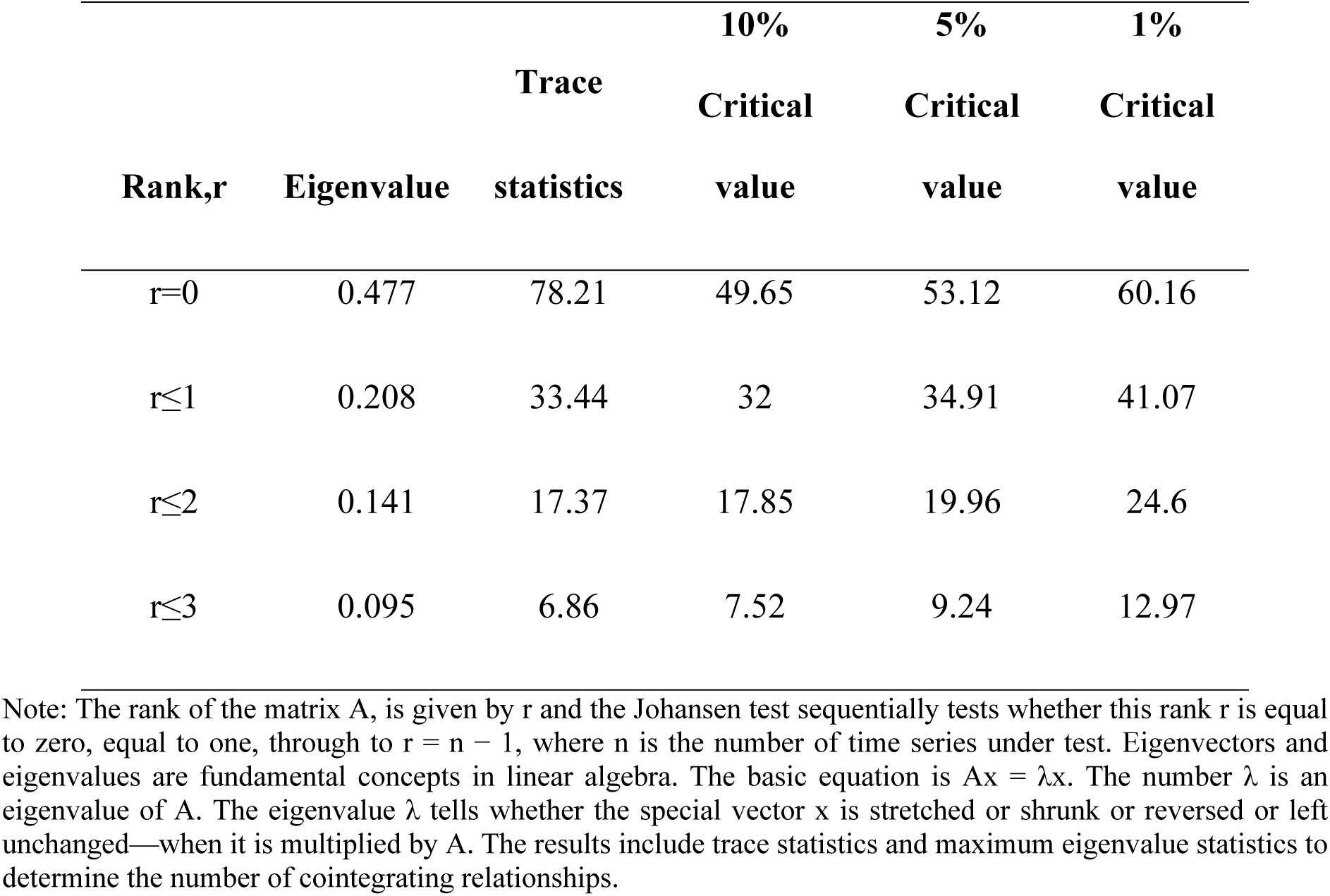
Johansen Cointegration Test Results.

#### Cointegration Vector

This vector equation describes the long-term relationship between variables through the process of cointegration of time series. The procedure of normalising each vector equation in Table 2 reveals the major dynamic relationship of cointegration. Over the long term, there is an inverse interaction between temperature and the sticky ovitrap index (SOI) with dengue cases, as compared to the ovitrap index (OI). This informative equilibrium provides us with the information that; persistence increment of temperature leads to decreases in dengue cases. The consistent and efficient sticky ovitraps, which capture adult mosquitoes over a lengthy period of time, have resulted in a decrease in dengue incidence. A consistently positive correlation between the coefficient of OI and dengue cases validates that OI can effectively serve as a reliable estimate of the aedes population in areas with a high burden of dengue.

**Table 2.** Cointegrating Vector Equilibrium.

| Variable | Coefficient |
| --- | --- |
| CASE | 1.0000 |
| TEMPERATURE | 0.9507 |
| OVITRAP, OI | 0.9566 |
| S. OVITRAP, SOI | 1.3755 |

#### VECM Estimation

When cointegration data series are present, the VECM model is used to calculate and include error correction terms that take into account any deviations from the long-term equilibrium. This model is well-suited for examining the short-term interactions between variables. A lag of p - 1 was included to adjust for the degrees of freedom used by the lag inclusion. Here is the VECM equation for this analysis:

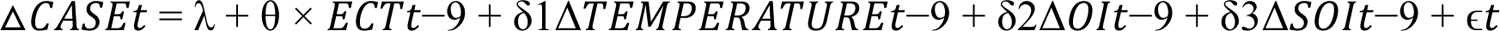

- Δ denotes the first difference of a variable.
- *ECT* _t-9_ (Error Correction Term) is the lagged value of the deviation from the
- long-term equilibrium (residuals from the cointegration equation), included to bring the variables back towards equilibrium.
- γ, θ, δ_1_, δ_2_, δ_3_ are parameters to be estimated.
- ɛ_t_ is the error term.

The model estimation requires the determination of 152 slope parameters, which illustrates the intricate and strong nature of the model in accurately representing the changes over various time intervals and interactions. The Akaike Information Criterion (AIC) and the Bayesian Information Criterion (BIC) quantify the goodness of fit and complexity of the model. The AIC has a value of 869.1324, while the BIC has a value of 1213.156. These criteria are useful in evaluating the model’s efficacy which is the trade-off between accuracy and complexity. This document elucidates the manner in which the variables being examined establish a connection within the equation. A model may exhibit exceptional fit or accuracy, although it lacks the capacity to elucidate the intricacies of variables. The greater the complexity of the dynamic, the lower the fit. In addition, the sum of squared residuals (SSR) is reported as 5510.52. This value serves as the total squared deviations of observed values from their predicted values within the model framework also known as a measure of the model’s overall error magnitude. Furthermore, it can be inferred that the model estimation has the capacity to elucidate 74.2% of the dynamics.

The VECM analysis shown in Table 3 offers a comprehensive understanding of the short-term interactions among dengue cases, temperature, and mosquito indices (ovitrap and sticky ovitrap) throughout time. The results reveal significant coefficients, especially in the error correction term and several lagged variables, demonstrating the impact of previous values on current trends in the data. In the case equation, the error correction component (ECT) has a substantial negative coefficient (−0.2584). It indicates deviation from the long-term equilibrium, will be corrected around 25.84% in each period. Therefore, the system has a strong tendency to regain back to stability. This highlights the efficacy of the dynamic interaction which enable it to revert to its equilibrium within the model estimated. The analysis further elucidates the dynamics of the significant variables;

i. The positive coefficients of sticky ovitrap index, SOI, in the sticky ovitrap equation at lags −6 and −8 demonstrated the sustained effectiveness of sticky traps. This indicates the crucial use of these tools as part of strategy in persistently high populations of the Aedes mosquito. This discovery suggests that the current existence of adult Aedes mosquitoes will have a substantial impact on the population’s growth after 6 and 8 biweekly intervals, corresponding to 3 and 4 months, respectively. However, at a lag of −1 (a gap of 14 days) and a negative coefficient, a low current capture measurement of SOI is associated with a past high SOI reading. This explains the ecological dynamics of Aedes mosquitoes and their life cycle, which includes the duration of the incubation period of the dengue virus. The dengue virus undergoes an incubation period of around 7 to 10 days inside the mosquito following its ingestion of blood from an infected individual. This explains that the particular phase of the life cycle is very much related to temporal fluctuations of the mosquito population and the impact on the utilization of sticky ovitraps within the ecological system.
ii. The case equation shows significant positive coefficients at case lag −3, lag −7 and lag −8. It implies that preceding dengue cases have a continuing impact on future incidences. This allows us to understand that the transmission of dengue follows a cyclical pattern in the long term. Additionally, it is affected by previous outbreaks, which cause deviations or fluctuations from a stable state. Consequently, this results in peaks in dengue cases or epidemics.
iii. In the case equation, in relation to SO variable at lag-3 and −4, conversely significant negative coefficients reveal that effective aedes control underscoring the effectiveness of these tool as interventions over time.
iv. The ovitrap equation in relation to ovitrap variables at lag-1, −3, and −4 produces negative coefficients, suggesting that a persistent increase in aedes larvae leads to a subsequent decline of the population in a time frame of two weeks up to two months. It indicates that there’s a rapid (brief period of time) external influence resulting in the decline of Aedes larvae. This might as well be the result of prompt control measure or interplay of other factors (latency −1, −3, −4). Nevertheless, the population of larvae will ultimately increase as a result of the inability to maintain control measures or other interactions that arise within the ecological system at lag-9, or 4.5 months later.
v. The significant negative coefficient of temperature at lag-6 for *case equation,* shows the interplay of persistent increasing of temperature in the ecological link led to decline of dengue cases subsequently.
vi. In the equation for the sticky ovitrap, the lag-1 case variable has a positive coefficient. Within a 14-day period, an increase in adult aedes mosquito populations will result in the transmission of dengue fever and the development of cases.

**Table 3.**
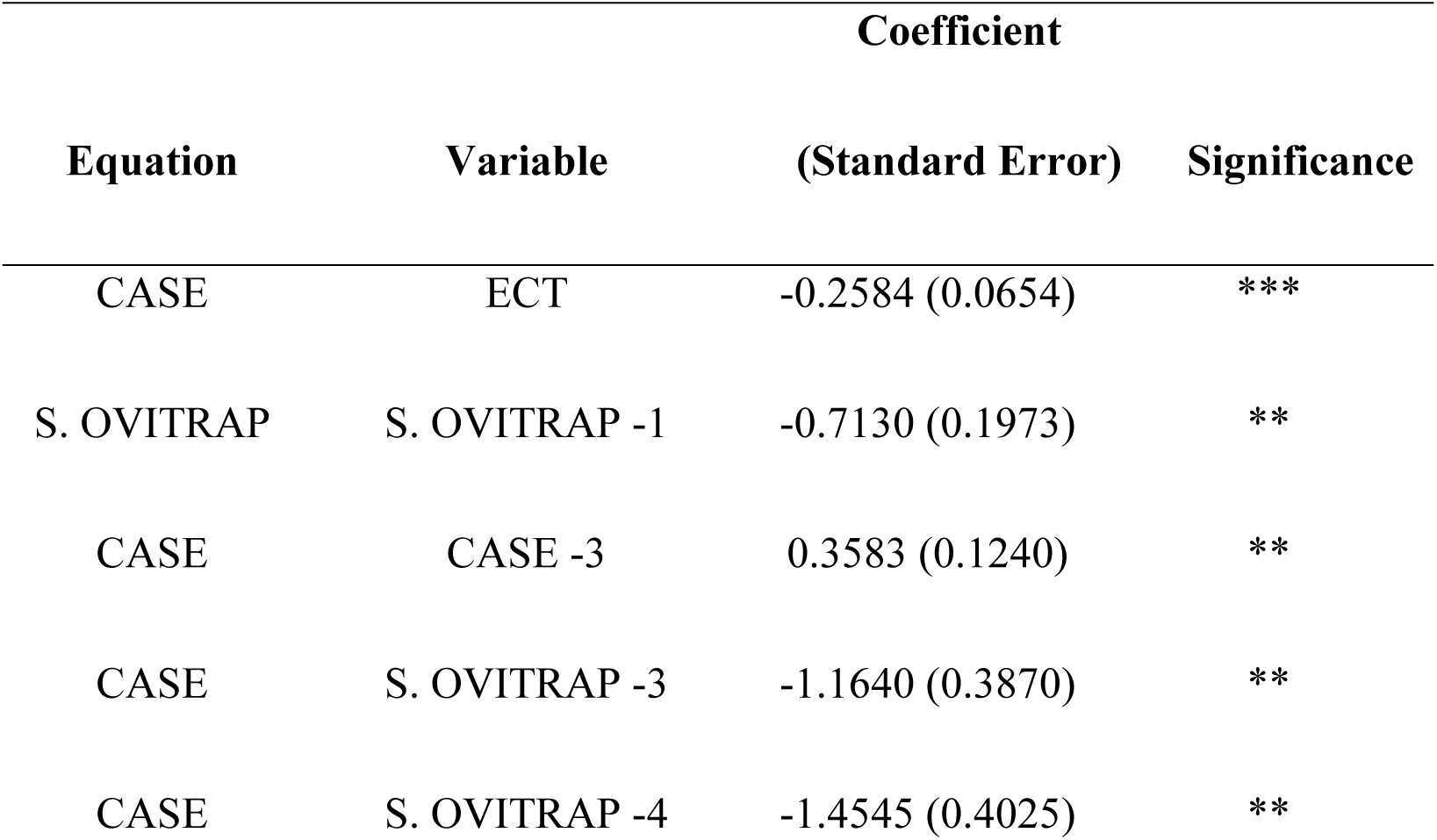
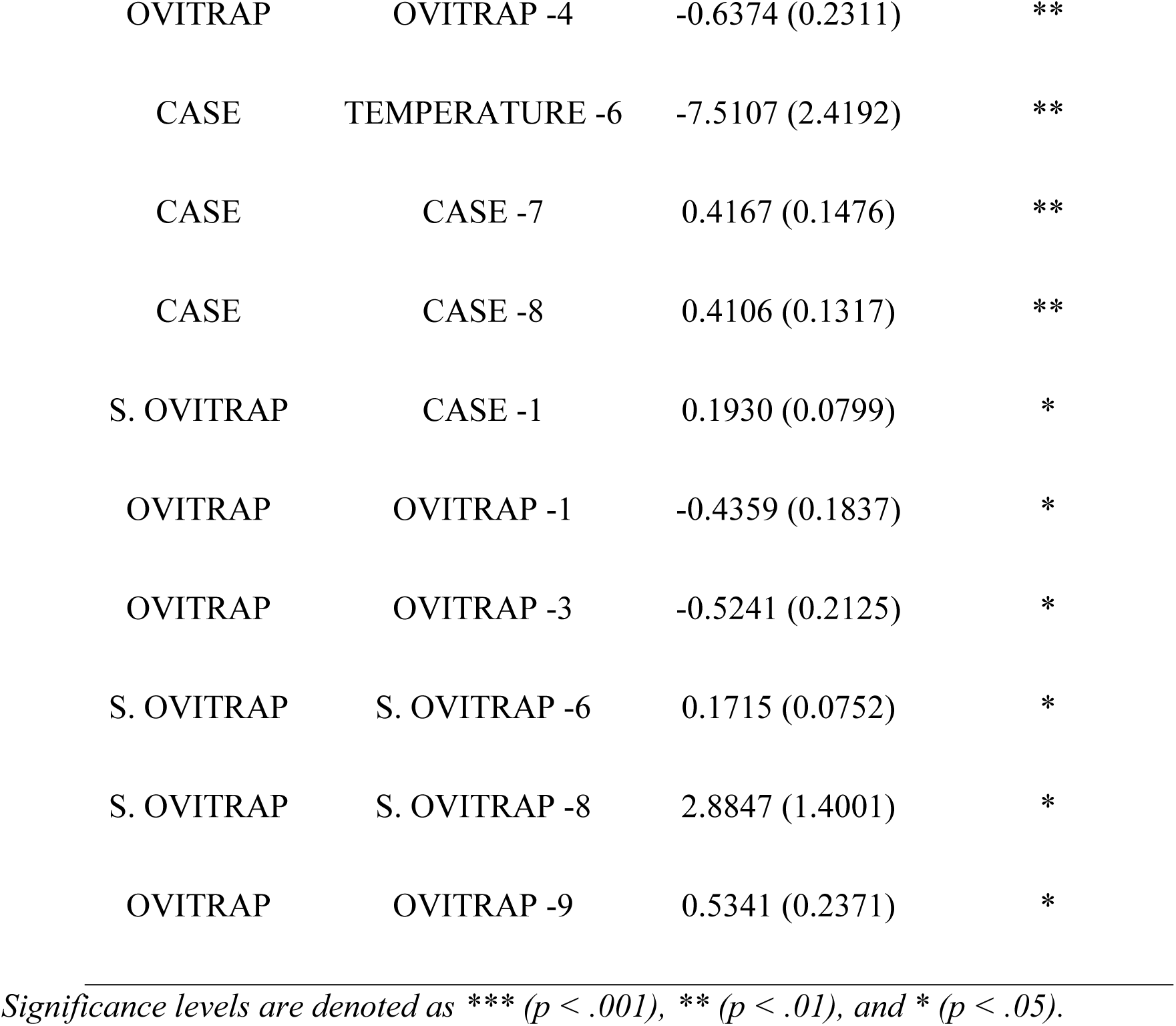
Significant Coefficients in VECM Analysis.

#### Forecasting based on VECM

In the evaluation of the Vector Error Correction Model (VECM) for forecasting purposes, the forecasted biweekly onset values were meticulously compared against the biweekly average actual values observed over a 12-week period, commencing from January 2023 until March 2023 (Figure 2). This comparative analysis aimed to assess the model’s predictive accuracy (Table 5) and reliability in estimating real-world occurrences within the specified timeframe.

**Figure 2.**
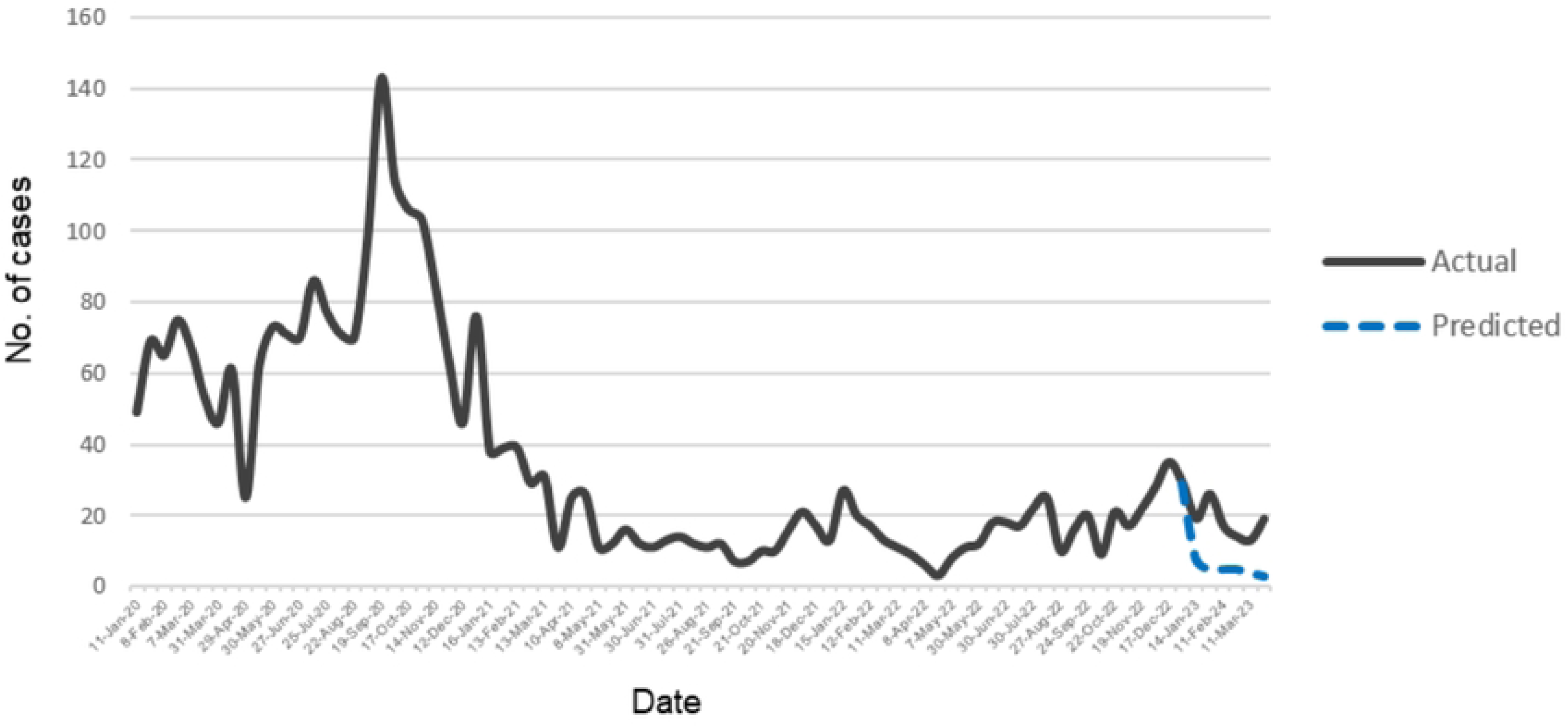
Actual case count line graph from January 2020 to March 2023 and predicted case count line graph of dengue case plotted from January 2023 to March 2023, extracted from R.

**Table 5.** Statistical metric values.

| Metric | Value |
| --- | --- |
| MSE | 176.2 |
| RMSE | 13.28 |
| MAPE | 2.72% |

The two main statistical metrics used to measure the differences between the VECM predictions and the actual observed values were the mean squared error (MSE) and the root mean square error (RMSE). The mean squared error (MSE) between the actual and predicted values is 176.42, which indicates the variance in forecast accuracy. While the RMSE, computed as 13.28 is directly relates to the average error magnitude. Additionally, a third metric known as the mean absolute percentage error (MAPE), was determined to be 2.72%. This sums a much clearer and easily interpretable indication in percentage terms. This metric is useful in delivering the relative accuracy of the forecasts, allowing stakeholders to look into the potential impact of forecast errors in operational terms [18,19].

The model is tested for this particular period of time, spanning from January to March of 2023. It matters for several reasons. First off, the study period was elongated to investigate a wider range of environmental and potentially epidemiological dynamics relevant to the topic of the research. These interplays included temperature deviations, vector populations as expressed by the ovitrap index (OI), and the sticky ovitrap index (SOI). It captures their effects on dengue fever incidences. Second, by examining model performance under different settings, data from this particular timeframe may be analyzed. This can reveal information on the adaptability and sensitivity of the VECM to temporal changes in the underlying variables.

## Discussion

This VECM method is distinct in its ability to establish both the long-term and short-term deviations of variables, including those related to epidemiology, entomology, and meteorology within an ecological context. It is a multivariate regression time series analysis that reduces cofounding factors, demonstrates variable cointegration, captures the causal effect either directly or inversely, and can be utilized for further forecasting. Knowledge of all the variables studied is essential to estimating the best fit and yet being able to elucidate the complexity at the same point. These findings are instrumental for the field workers, public health officials and policymakers, emphasizing the need for historical data consideration and continuous analysis and the implementation of timely, sustained interventions to manage and mitigate dengue effectively. The key findings are further described;

### Temperature

This study showed that the continued rise in temperatures results in a decline in the number of dengue cases over an extended period of time. Additionally, it recorded a temporary divergence, referring to a variation in temperature from its stable state that will influence dengue instances three months later. Nevertheless, the temperature has shown a zero-lag association with sticky ovitrap and ovitrap index. The dynamic of the interaction between temperature and dengue cases can be attributed to the ecological system, namely the complicated impact of temperature on vector transmission to humans.

### Temperature effect on biological characteristics of aedes

Mosquitoes are poikilothermic organisms, meaning that variations in the ambient temperature have an immediate impact on their internal body temperature. Although warmer temperatures can initially facilitate mosquito breeding and virus transmission, they can also result in long-term ecological adaptations that help control mosquito populations. These adaptations may include greater predator activity or faster virus incubation, which can lead to early viral die-offs [20]. The ideal temperature range for their growth and progress is between 25 and 30 degrees Celsius. As the ambient temperature increases, there is a proportional increase in the activity of Aedes mosquitoes [21]. Studies have demonstrated that Aedes albopictus displayed the shortest gonotrophic cycle at a temperature of 30 °C, which is within a duration of 3.5 days [22]. Carrington et al. observed that the duration of the gonotrophic cycle in Aedes aegypti decreased as the average temperature increased [23]. Thus, evidence has shown that temperatures greater than the ideal range can result in a decrease in lifespan and deter immature stages development. This will result in a significant reduction in the number of eggs produced, which leads to a decrease in the size of the population [24–26].

### Temperature effect on vector competence

Vector competence refers to the ability of mosquitoes to contract infections and then transmit those infections to other hosts. The mechanism remains incompletely comprehended. Generally, Ae. albopictus exhibits lesser vector competence compared to Ae. aegypti. The transmission of the dengue virus (DENV) was enhanced as the temperature climbed within the range of 18°C to 32°C. Nevertheless, the transmission of the virus is diminished when the temperature exceeds 32 °C, resulting in a decrease in its ability to be transmitted by its carrier [27,28]. This is intimately related to immune system pathways and tissue barriers in the mosquito, which lower the rate of midgut infections and DEN V transmission [29].

### Temperature effect on Den Virus, DEN V

DEN V is a single-positive-stranded RNA virus that encodes three structural proteins and seven non-structural proteins. The structural proteins consist of the capsid (C), membrane (M), and envelope (E) proteins. The temperature-induced structural alterations usually occur between 31°C and 35°C and are irreversible, weakening the E protein [30,31]. Hence, it diminishes viral infectivity. Therefore, it is imperative to thoroughly examine the impact of global warming on the survival of Aedes mosquitoes. This will certainly interfere with the transmission of dengue fever. The impact on warmer climate regions results in being closer to the thermal thresholds that many different kinds of species have. It is also necessary to consider the impact on cooler regions, which encourages the spread of vector borne illnesses [32]. Consequently, the burden and rate of transmission of vector borne diseases are significantly impacted by this modification. Therefore, it is essential to closely monitor this climate change signal and how it affects the adaptability of the vector.

### Sticky Ovitrap

In the context of epidemics, the conventional approach of employing insecticides to manage adult female aedes mosquitoes has been replaced by a new approach that focuses on the creation and evaluation of non-insecticidal methods. Moreover, the increasing apprehension regarding the development of pesticide resistance and its adverse effects on the environment and human health is a significant catalyst for the adoption of safer and more sustainable technologies. The sticky ovitrap has become increasingly significant in the monitoring and management of mosquitoes in recent years [33,34]. It offers a quicker and more direct assessment of the efficacy of ovipositional attractants, while also catching the carrier of circulating Den V. There is a scarcity of research on the use of time series or regression analysis to the use of sticky ovitraps. The findings of this study demonstrate that consistent deployment of sticky ovitraps in an area with a high prevalence of dengue fever has proven to have a direct impact on the development of dengue cases. It showcased its utility and efficacy as a component of a control approach.

The short-term sticky ovitrap equation, coupled with the sticky ovitrap variable, illustrates the interplay between the bionomics and life cycle of Aedes at lag 1. Furthermore, it indicates that the ongoing existence of adult aedes mosquitoes will dictate the population’s capacity to thrive approximately 3 to 4 months later. Thus, incorporating it into the control plan in areas with consistently high dengue cases or outbreaks proved to be an invaluable tool. The intervention tool becomes significantly more successful over time, namely at 1.5 and 2 months later. However, when considering a short time period (specifically, a lag-1 or 14-day interval), the interaction between the sticky ovitrap equation and the case variable showed that the tool acts primarily as a surveillance tool rather than a control tool. This suggests that there is a possibility for the rapid and continuous transmission of the dengue virus to result in the occurrence of new cases. Therefore, in an area where there is a significant presence of adult aedes mosquitoes, it is crucial to implement prompt and intensive control methods in conjunction with the use of sticky traps. It also implies subsequent measures or evaluations in the area affected by dengue is crucial after implementing an initial control intervention or first control cycle.

Recent studies have demonstrated encouraging effectiveness in utilizing mass trapping of lethal ovitraps to control the aedes population. Nevertheless, the level of certainty regarding these findings is only moderate. Several key elements are missing, such as the study design, the operationalization, the standard index used, and the analysis of the data, including the outcome or epidemiological endpoint [33]. The outcomes of this model’s dynamic analysis will unequivocally demonstrate the efficacy of lethal ovitrap intervention in complementing measures to mitigate dengue cases.

Two types of Aedes adult traps can be deployed depending on the physiological stage of the targeted Aedes mosquitoes: (i) host-seeking female traps and (ii) lethal ovitrap gravid females. Many countries have used these traps as part of trial interventions [34]. To achieve a significant reduction in mosquito populations and viral transmission, it is important to study the protocols for operational deployment circumstances in order to identify the best deployment parameters under various epidemiological scenarios. It is imperative to take into account the guidelines of the World Health Organization and adapt them to the specific operational conditions of the control team [35]. The extensive utilization of deadly ovitraps as a component of the dengue control strategy provides substantial support for the 3rd and 11th Sustainable Development Goals (SDGs). Addressing a logistical and financial investment setback in the trap’s deployment is crucial. In conjunction with that, cost-effectiveness and cost-benefit should be carried out as part of an economic evaluation measure.

### Ovitrap

The long-term equilibrium indicates that the ovitrap tool can be used as a reliable predictor of dengue cases in an area with a significant dengue burden. Consistently high current OI readings in an area will determine the persistence of the larval population between four and five months later. This marker can certainly serve as an invaluable tool to help in informed decision-making for public health officers and stakeholders. This decision consists of either continuing with the use of ovitrap as a monitoring tool that can trigger appropriate control measures or using it as a marker for exit strategy after a certain time frame prior to relocating the tool to another site. A recent forecasting study conducted in Brazil, employing a different analytical model, corroborates this claim by indicating that the utilization of ovitraps has the potential to identify dengue epidemics with a lead time of four to six weeks [36].

### Dengue case

This analysis expressed the temporal pattern of dengue transmission, which is persistent and cyclical in nature while, concurrently, in the short term, being influenced by past outbreaks, giving rise to deviations from equilibrium, thus resulting in peaks of dengue cases or epidemics. Hence, this research offers a valid agreement for the execution of a successful dengue epidemic plan. Efficient management of outbreaks can effectively avert the occurrence of explosive and rapid spread of epidemics in the future.

Intriguingly, many countries in Southeast Asia exhibit a cyclical trend, experiencing surges in dengue infections throughout specific periods. The apparent high prevalence of dengue in the Southeast Asia region can be partly attributed to hyperendemicity, which refers to the simultaneous circulation of all four serotypes of the virus [36]. The occurrence of recurring epidemics involves the rotation of DEN V serotypes. Empirical evidence has shown that the variation in the yearly dengue pattern is caused by the interchanging of serotypes or the displacement of serotypes [37,38]. Counterfactual study suggests that the presence of dengue hyperendemicity greatly amplifies the intensity of prior outbreaks [39]. It corresponds to this model of interaction dynamics, which elucidates the surge in the number of cases in the current outbreak and is connected to prior dengue occurrences. Furthermore, the intricate balance is influenced by vector dynamics and immunological interactions [40,41].

### Limitation

This study does not consider variables such as demography, population immunity, mosquito virus surveillance, or the socio-economic structure of society. Historically, there has been a lack of consistent collection of serial data on virus surveillance in mosquitoes. This study also recognizes the significance of vector control in the potential disruption of equilibrium. This study focused exclusively on dengue cases that were diagnosed and reported solely to the health district. Hence, the system’s ability to accurately document the overall number of dengue cases may be compromised due to the presence of diverse dengue symptoms, resulting in instances where illnesses go unnoticed or undetected.

### Future research

In light of the preceding findings, it is paramount to develop a certain standard of intervention index to address the inadequacy of consistency of method when assessing various interventions. It will be tremendously useful and convenient for evaluating and comparing the various measures that have been adopted in combating dengue in a temporal manner. The deployment of sticky ovitrap as a mass trapping control method under sustainable engagement circumstances could later be studied and explored with the adaptation of control protocols. Temperature is indeed an overall excellent estimator of the meteorological parameter that has a direct impact on the development of dengue cases. It is recommended to use more extensive data to look into its temporal interaction with ovitrap, sticky ovitrap findings, and progression of virus in mosquitoes.

## Conclusions

The VECM model provides an excellent trade-off between goodness of fit and complexity in describing the variables examined. Mean absolute percentage error (MAPE), computed as 2.72%, is an easy interpretation and best for describing the forecasting performance. The careful juxtaposition of the prediction values with the actual values that were observed during the first quarter of 2023 underscores the challenges relevant to a real-world phenomenon. Previous dengue occurrences predicted a surge of new dengue cases while preserving the cyclical pattern. The model additionally predicts the utility and efficacy of sticky ovitrap, thereby establishing and reinforcing its dual roles in aiding strategy. The result also reinforced the use of ovitrap as a surveillance tool and offered substantiation of the influence of temperature on the progression of dengue cases. Refining these results into strategies will ultimately enhance public health responses to vector-borne diseases. The magnitude of quantified differences illustrates the intricate interactions of determinants that influence dengue transmission. Despite these limitations in modelling, the VECM is able to approximate the long-term equilibrium and short-term adjustments between the variables, which allows it to be exploited for progressively informing public health policies and strategies.

## Data Availability

: All data is available in the manuscript and the figures

## Acknowledgments

The authors are grateful for the Ministry of Health Malaysia.

